# HbA1C and Fructosamine levels during Infancy

**DOI:** 10.1101/2025.05.07.25327198

**Authors:** Rebecca Schneider Aguirre, Abeer Al-Raddadi, Linda A. DiMeglio, Erica A. Eugster

**Author notes:** Corresponding Author: Rebecca Schneider Aguirre. RSA is currently at Baylor College of Medicine. AA-R is currently at Michigan State University, Lansing. The collection and analysis of the current data occurred at Indiana University. <u>Author Email addresses:</u> Abeer Al-Raddadi, Linda A. DiMeglio, Erica A. Eugster.

## Abstract

Due to a high percentage of hemoglobin F, Hemoglobin A1C (HbA1C) measurements are inaccurate for assessing glycemia in infants. We aimed to determine when HbA1C might have utility and to assess the value of fructosamine. We measured HbA1C in healthy infants aged 3 weeks to 12 months. Hemoglobin, HbA1C, hemoglobin electrophoresis, fructosamine, and albumin levels were collected. Mean age was 193.9 ± 94.5 days; participants were 60.9% male, 80.4% white, and 15.2% Hispanic. Mean HbA1C (n=31) and fructosamine (n=33) were 5.0% (31 mmol/mol) (range 4.4–5.9%; 25–41 mmol/mol) and 217 (range 186–261 mCmol/L), respectively. HbA1C percentages negatively correlated with HbF percentages (p < 0.005) and rose with increasing age in infants <6 months (p < 0.01). Fructosamine did not vary with age. Normalizing HbA1C to hemoglobin fractions or fructosamine to albumin did not change significance. We conclude that HbA1C values (via HPLC) likely become a reliable marker of glycemia after 6 months of age and that fructosamine may be a better measure during this young age.

## Introduction

The risk of long-term complications associated with diabetes is related both to glycemia, historically measured via glycated hemoglobin (HbA1C), and duration of diabetes. The American Diabetes Association 2024 Standards of Care identifies HbA1C values as the “primary tool for assessing glycemic status” due to the linkage with complications^1^. However, the reference HbA1C needed to reduce the incidence of complications was developed based on data in an adult population^2,3^. Infants have high concentrations of fetal hemoglobin F (HbF) and experience high erythrocyte turnover as the HbF to adult hemoglobin transition occurs^4,5^, thereby impacting HbA1C levels. Infants with neonatal diabetes, defined as diabetes diagnosed within the first 6 months of life, require serial monitoring of average glycemia and, in some cases, insulin treatment^6^. Those with neonatal diabetes also have the potential for longest duration of disease and limited data exist regarding HbA1C measurements in infants. The American Diabetes Association has recognized other glycemic measures, including fructosamine and glycated albumin, in persons for whom HbA1C may not be reliable^1^. While small studies have examined the use of fructosamine in infants^7–10^, none has examined if an age-dependent reference range exists nor if fructosamine and HbA1C levels correlate in young children. Continuous glucose monitoring (CGM) is becoming more and more utilized as a modality for assessing glycemic control. However, while some studies have shown correlation between time-in-range and estimated A1C^11^, the landmark studies demonstrating reduced long-term complications utilized HbA1C values^12–15^. Furthermore, 2 years is the youngest age of CGM FDA approval. Thus, no standard of care measurement of glycemic control has been characterized in this age-group.

Due to the American Diabetes Association’s emphasis on HbA1C as the “primary tool for assessing glycemic status” and recognition of alternative measures such as fructosamine, we obtained HbA1C and fructosamine levels in infants without diabetes to assess the utility of both measures in this age group.

## Research Design and Methods

This study was approved by the Institutional Review Board at Indiana University and conducted between November 2019 and June 2021 at Riley Hospital for Children at Indiana University (IU) School of Medicine, Indianapolis, IN. Participants were aged 3 weeks through 12 months, without risk factors for dysglycemia, and undergoing venipuncture for non-study (clinical care) reasons (see Supplemental Tables 1 and 2 for exclusion/inclusion criteria and venipuncture reasons, respectively^16^). Written informed consent was obtained from a parent or guardian. The study was designed for prospective enrollment; the study design was amended to also include laboratory remnant samples when recruitment slowed during the spring of 2020 due to the pandemic. Medical history was obtained from chart review and confirmed by participant report for prospective enrollees.

Hemoglobin, hemoglobin electrophoresis, HbA1C (via high-performance liquid chromatography or HPLC), and albumin levels were measured at IU Health Hospital Laboratory, Indianapolis IN. HPLC separates the different hemoglobin fractions into a chromatogram with different peaks, which are used to measure the percental of HbA1C. The HbF peak is next to the HbA1C peak. Fructosamine was measured at ARUP Laboratories (via quantitative spectrophotometry). Due to difficulty obtaining a sample or hemolysis, participants may not have results for every category.

### Statistical analysis

HbA1C was normalized by dividing HbA1C percentage by either HbA, HbA2 or HbF percentages. To correct for HbF percentages, HbA1C was calculated as a percentage of total Hb minus HbF. Fructosamine was normalized by dividing the fructosamine amount by the total albumin amount. SPSS (Version 24, IBM Chicago, IL) was used for all statistical calculations. p values were calculated with a level of <0.05 accepted as significant.

## Results

Participants were 60.9% male, 80.4% white, and 15.2% Hispanic (Table 1); mean age was 193.9 ± 94.5 days (range 22 - 363 days) (Table 2). Supplemental Table 3 compares racial demographics of study and surrounding populations^16^. Children had venipuncture related to sedation (n=26) or need for labs (n=20). Congenital hypothyroidism (n=4) was the most common reason for lab draws. Almost half (48.6%) of all enrolled infants used at least one medication; Vitamin D was the most common (n=10), followed by levothyroxine (n=4). Supplemental tables 2 and 4 list the conditions evaluated and medications used, respectively^16^.

**Table 1:** Demographics of Enrolled Participants (n=46). Gestational age, birth data, racial and ethnic information were based upon self-report from the parents and guardians or chart review (for remnant samples).

|  |  |
| --- | --- |
| **Mean Age:** | 194 ± 95 days |
| **Sex:** | 60.9% male |
| **Race:** | 80.4% white, 6.5% black, 2.2% Asian, 8.7% biracial, 2.2% triracial |
| **Hispanic Status:** | 15.2% Hispanic |
| **Mean Weight:** | 7.6 ± 2.00 kg |
| **Mean Length:** | 65.8 ± 6.2 cm |
| **Mean Birth Weight:** | 3.35 ± 0.44 kg |
| **Mean Gestational Age:** | 38.6 ± 1.8 weeks |
| **On any medication?:** | 48.6% yes |
| **Reason for venipuncture** | 56.5% due to sedation; 43.5% medical evaluation |

**Table 2:** Mean A1C and Fructosamine Values by Age Group. Samples are grouped according to the participant’s age at the time of the blood sample. The samples in parentheses indicate the number of resulted samples, first number for the HbA1C group and second number for the fructosamine group. HbA1C values could not be obtained from samples in 9 participants due to either high HbF levels (n=7, range HbF 16.4-85.4%), high amount of hemoglobin variants (n=1), or insufficient sample (n=1). One participant had an HbA1C percentage outside the reference range at 5.9%. This participant had no risk factors nor symptoms of hyperglycemia and had a normal fructosamine level. One-way ANOVAs were done between age groups for both HbA1C and fructosamine; fructosamine means are not significantly different but HbA1C means are significantly different (p<0.02).

| **Age groups** | **N**<br>**(samples with results)** | **HbA1C mean**<br>**Ref. range: 4.0-5.6%;**<br>**20-38 mmol/L**<br>**(range)** | **Fructosamine mean**<br>**Ref. range: 170-285**<br>**mCmol/L**<br>**(range)** |
| --- | --- | --- | --- |
| **0-2 months** | 7<br>(0, 4) | none | 218<br>(186 – 239) |
| **3-4 months** | 8<br>(6, 7) | 4.8%; 29 mmol/L<br>(4.4 – 5.0; 25-31) | 215<br>(191-261) |
| **5- 6 months** | 7<br>(6, 6) | 5.1%; 32 mmol/L<br>(4.5 - 5.5; 26-37) | 222<br>(188-256) |
| **7- 8 months** | 11<br>(10, 10) | 4.9%; 30 mmol/L<br>(4.4 – 5.2; 25-33) | 221<br>(194 – 250) |
| **9-10 months** | 4<br>(4, 4) | 5.5%; 37 mmol/L<br>(5.1 – 5.9; 32-41) | 215<br>(206 – 228) |
| **11-12 months** | 5<br>(5, 2) | 5.1%; 32 mmol/L<br>(4.8 – 5.3; 29-34) | 206<br>(199-212) |
| **Total** | 42<br>(31, 33) | 5.0%; 31 mmol/L<br>(4.4-5.9; 25-41) | 218<br>(186-261) |

A flow diagram of enrolled infants is presented in Figure 1.

**Figure 1.**
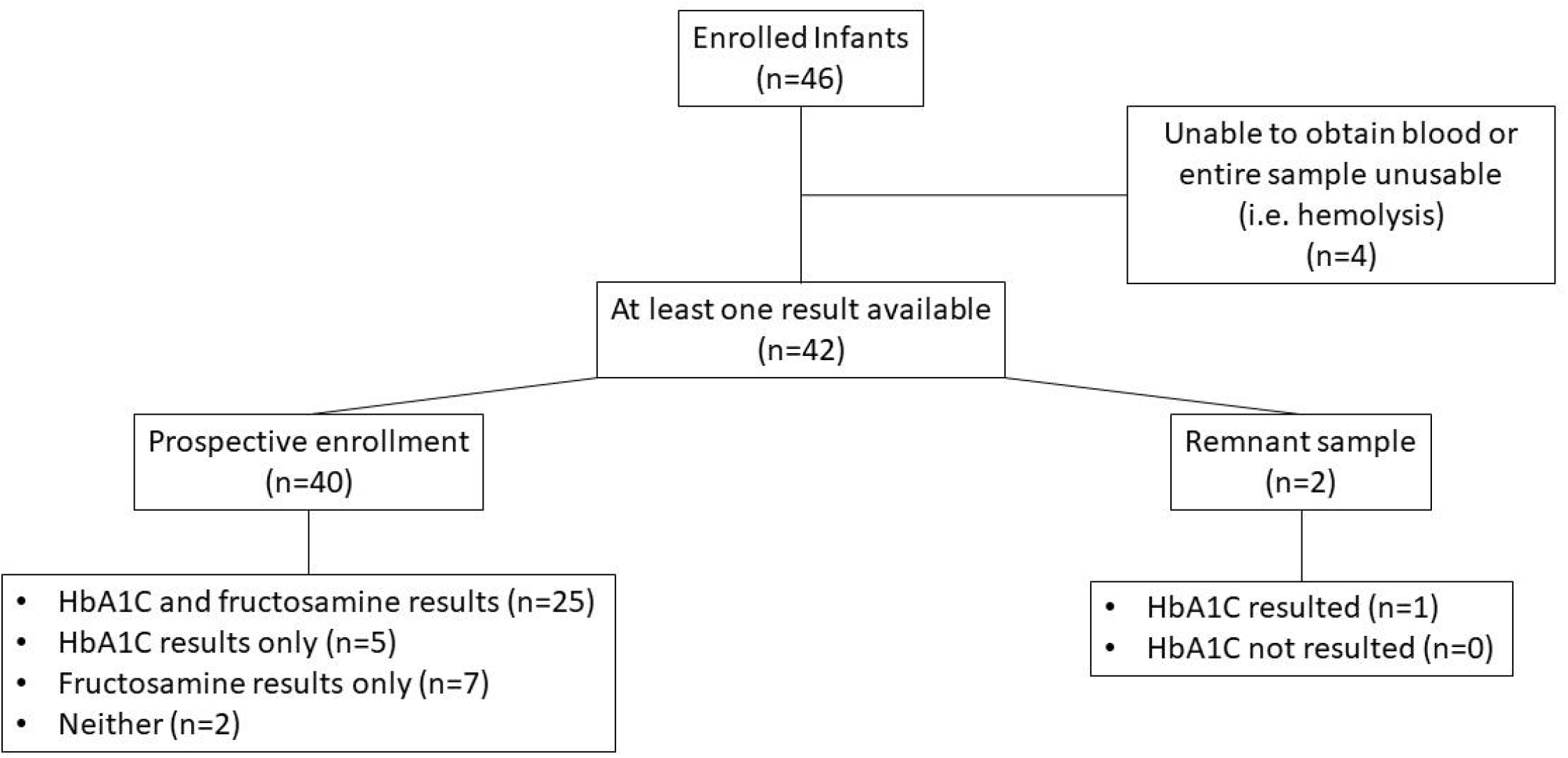
Enrolled Infants and Resulted Samples. Flow diagram of all enrolled infants. In four infants, either blood was unable to be obtained or it was of insufficient quantity or quality for the assay. In infants in which blood was obtained, but either A1C was not resulted (remnant sample) or neither HbA1C nor fructosamine was obtained (prospective enrollment), hemoglobin electrophoresis was able to be performed (often with a high percentage of HbF) and thus these samples were included for analysis. Due to assay requirements, only HbA1C and Hb values were obtained in remnant samples. *Hemoglobin and hemoglobin electrophoresis results were available.

HbA1C levels were obtained in 31 participants, with a mean HbA1C of 5.0% (31 mmol/mol). Fructosamine levels were obtained in 33 infants, with a mean fructosamine of 217 mCmol/L. We grouped HbA1C and fructosamine levels according to infant age by 2-month increments, with the means and ranges presented for each group (Table 2). HbF levels ranging from 16.4 – 85.4% interfered with obtaining an HbA1C percentage; these infants were aged 47 – 255 days (median age, 84 days). The youngest age a HbA1C value could be obtained was 122 days. There was an overall trend toward a positive correlation of age and HbA1C (p < 0.056) (Fig. 2A). This was driven by results in infants younger than 6 months, where HbA1C values positively correlated with age (p < 0.01) (Fig. 2B). No correlation was present for infants older than 6 months (data not shown). HbF percentages, range 0.4 – 85.4%, were highly variable and decreased with older ages (p < 0.001; data not shown). Higher HbF was associated with lower HbA1C percentages (p < 0.005) (Fig. 2C). The highest HbF percentage for which an HbA1C value could be obtained was 12.4%. No significant differences existed when HbA1C was normalized to HbA, HbA2 or HbF (data not shown). Since uncontrolled hypothyroidism is associated with anemia, we examined the total Hgb levels in hypothyroid infants compared to study age-group and none of the infants had the lowest level for age group (Supplemental Table 5)^16^. All infants except one had free T4 levels above the lower limit of normal; the one infant with a low free T4 level did not have a resulted HbA1C percentage (Supplemental Table 5)^16^.

**Figure 2.**
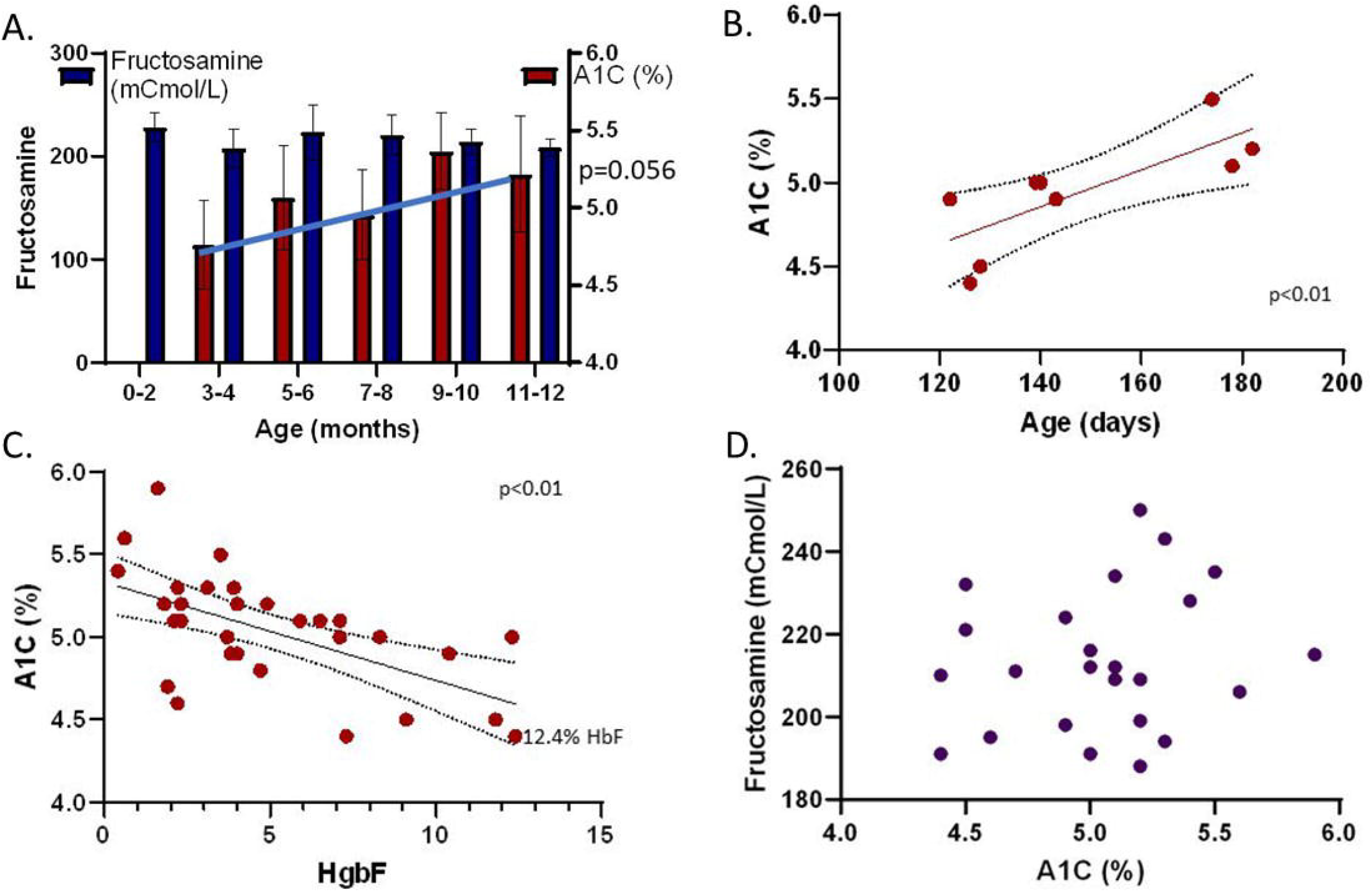
Fructosamine and HbA1C levels by age and HbF. A. Average Fructosamine and HbA1C levels by age. All available HbA1C samples were used to calculate the correlation between age and HbA1C levels. Results are shown as means ± SD. The youngest age for which an HbA1C value was obtained was 122 days. A correlation between age and HbA1C level trended towards significance (p<0.056). There was no correlation between age and fructosamine levels. [n for A1C values by month age group were: 0-2, 0; 3-4, 5; 5-6, 6; 7-8, 11; 9-10, 3; 11-12, 6. n for fructosamine value by month age group were: 0-2, 3; 3-4, 7; 5-6, 8; 7-8, 10; 9-10, 3; 11-12, 3.] **B. HbA1C levels increase with age in those younger than 6 months old**. A separate correlation (than in A) was calculated for infants ≤182 days (n=9). p<0.01. **C. A1C levels decrease as HbF levels increase**. p<0.01. HbF levels from all participants ranged from 0.4% to 85.4%. The highest HbF level for which a HbA1C level was obtained was 12.4% (n=31). **D. Fructosamine levels do not correlate with HbA1C levels**. The correlation coefficient was 0.18 and p = 0.39. (n=25, samples with both HbA1C and fructosamine.)

All fructosamine values were within the reference range. In contrast to HbA1C, they did not correlate with age (Fig. 2A). Normalizing fructosamine to albumin did not change this observation. Fructosamine levels did not correlate with HbA1C levels (Fig. 2D).

HbA1C and fructosamine levels did not differ by sex, race, ethnicity, medication use, gestational age, or current weight (data not shown).

## Discussion

Here, we characterized HbA1C and fructosamine levels in infants, showing that HbA1C obtained by a HPLC method is not measurable until HbF values are ≤12.5%, that HbA1C levels positively correlate with age in those <6 months, and are negatively correlated with HbF levels. In contrast, fructosamine levels were all within the reference range, and do not correlate with HbA1C levels nor age.

Our study both correlates with results of previous studies and adds fundamental information to the existing literature. Previous studies of HbA1C levels in infants without diabetes have focused either on neonates or older infants aged at least 8 months^9,17–20^. The reported mean HbA1C values here are comparable with those studies. However, we obtained HbA1C levels cross-sectionally over a wider age range, from 3 weeks through 12 months, and present the HbA1C ranges according to 2-month age intervals as well as the entire cohort.

HbF levels are a significant determinant of HbA1C percentages in early infancy: both due to a relatively large quantity at this age and that the HbF γ chains are less prone to glycation than the HbA β chains^21^. In a prior report, HbA1C levels were obtained longitudinally in 5 infants with NDM^9^ and inversely correlated with HbF percentages. Our results from the 31 infants in our cohort with presumed normoglycemia are complementary in that HbA1C percentages rose with age in those <6 months old and similarly were inversely correlated with HbF amounts. There was an overall wider spread of HbA1C percentages for HbF percentages when the HbF percentage fell below 5% (Fig 2), suggesting that HbF has minimal effect on A1C levels in this range, confirming a prior report of the same HbF threshold^22^. While HbF amounts appear to be a principle factor influencing HbA1C percentages in this age, physiological decreases in erythropoiesis and erythrocyte life-span occur during the first several weeks of life and likely also impact HbA1C levels^7^. HbA1C levels likely become a reliable predictor of overall mean glycemia after age 6 months: prior to age 6 months, HbA1C levels significantly increase with increasing age, indicating that age (and, likely, the associated decreasing HbF percentage during this time of infancy) is a confounder. However, after 6 months of age, there were no observed correlations between HbA1C levels and age, suggesting that age no longer influences HbA1C levels in older infants.

Current guidelines recommend that HbA1C percentages should be measured by a National Glycohemoglobin Standardization Program in an accredited laboratory and traceable to the Diabetes Complications and Control Trial (DCCT) method^1,23^. HbA1C can be assessed by a variety of different assays. We used an HPLC method, which was the DCCT primary reference method^23^. However, with this assay in infants, the presence of high concentrations of HbF creates a peak that overlaps with the HbA1C peak, rendering it impossible to determine the percentage of HbA1C. Other methods include immunoassay, electrophoretic methods, and boronate affinity chromatography, which are all prone to interference from HbF^22^. Mass spectrometry is very accurate without significant interference from HbF but is cost-prohibitive for routine clinical practice^21,22^. Therefore, considering the balance of practicality for routine clinical care and approved standards, we chose to use HPLC for HbA1C measurements.

Most previous studies of fructosamine levels in children have not provided granular data from healthy infants. They have included comparisons of children (not infants) and adults or in reference ranges in which all children aged 0-3 years (n=33) were included as one group^7,8^. Uniquely, our study used infants grouped in 2-month intervals, as well as the entire cohort (n=33, Table 2). The mean fructosamine level in our study is comparable to those obtained in the other studies. We did not find significant differences in fructosamine values between the different age groups, suggesting that fructosamine levels and norms may be relatively stable across the age ranges.

We surprisingly found no correlation between fructosamine and HbA1C levels. Since all fructosamine values fell within the reference range and were similar to previously published values for young children, fructosamine is likely a reasonable measure of glycemia in infants. However, infancy is characterized by changes in erythrocyte size, MCHC, lifespan, and HbF to HbA conversion, all of which impact HbA1C values^24,25^. Furthermore, since HbA1C percentages correlate with age in those younger than 6 months, but fructosamine levels do not correlate with age, too many variables regarding HbA1C percentages exist for true correlation between HbA1C and fructosamine. We suspect that fructosamine levels do correlate with average glycemia, although a study utilizing continuous glucose monitoring (CGM) is needed to confirm this.

A strength of this study is the focus upon the entire first year of life rather than one specific time-point (i.e. cord blood, a particular month of life) or older age groups. The number of enrolled infants is robust compared to prior work. The participants were infants who needed venipuncture for reasons outside of this study. While this was done to yield a larger sample size and to obtain samples in an ethical manner from children who otherwise would not need to have venipuncture, it is also possible that our results may not be generalizable to all infants. We tried to reduce this possibility by excluding known factors of dysglycemia in infants. We approached all eligible infants without respect to sex, race, nor ethnicity with the goal of enrolling a participant demographic mirroring the area (Supp Table 3)^16^. Regardless, Asians and Blacks are a small proportion in our study. Since glucose-6-phosphate dehydrogenase deficiency is more prevalent in those not of European descent, it’s possible that the HbA1C ranges could be different in non-white populations^26^. Hemoglobinopathies have a similar effect and we excluded infants with any known hemoglobinopathies. It is also possible that the infants with congenital hypothyroidism could have anemia from hypothyroidism superimposed upon the physiological anemia of infancy^27^. However, this is unlikely to occur without hypothyroxinemia^27^ and all those with assessable HbA1C had adequate free T4 levels (Supplemental Table 5)^16^. Furthermore, the total hemoglobin levels of each infant were within the group hemoglobin range (Supplemental Table 5)^16^. Additionally, since our study was cross-sectional in design we cannot evaluate HbA1C and fructosamine trends over time. Considering the increasing value and use of CGM, this modality could be used in future studies to determine normative data. However, at this time, no CGM has been approved or validated for under 2 years of age in either the United States or Europe.

In conclusion, we show that HbA1C values obtained by HPLC are likely reliable markers of glycemic control after 6 months of age and that values cannot be obtained until the HbF percentage falls below 12.5%. HbF percentages ≤5% appear to minimally affect HbA1C levels. If concern exists regarding the accuracy of HbA1C in an infant, HbF percentage can be evaluated using hemoglobin electrophoresis. Fructosamine levels do not appear to be affected by age and may be a better clinical marker of glycemia in very young infants.

## Data Availability

Some or all datasets generated during and/or analyzed during the current study are not publicly available but are available from the corresponding author on reasonable request.

