## Supplemental for "HbA1C and Fructosamine levels during Infancy"

**Supplemental Table 1: Reasons for venipuncture.** Condition describes either reason for labs (initial evaluation or monitoring). In the case of MRI, venipuncture was needed for sedation, and labs were collected at that time. Any number in parentheses indicates that there was that number performed for the condition. The condition represents the reason for the study but is not necessarily the final diagnosis.

| Condition | N (%) |
| --- | --- |
| Ambiguous Genitalia | 1 (2.2) |
| Bladder Extrophy | 1 (2.2) |
| Body Odor | 1 (2.2) |
| Congenital hypothyroidism | 4 (8.7) |
| Elevated TSH | 1 (2.2) |
| Failure to thrive | 2 (4.3) |
| Possible X-linked Hypophosphatemia | 2 (4.3) |
| Hypercalcemia | 2 (4.3) |
| Hypocalcemia | 1 (2.2) |
| Hypogonadism | 1 (2.2) |
| Osteopenia | 1 (2.2) |
| Pseudohypoaldosteronism | 1 (2.2) |
| Pubic Hair | 1 (2.2) |
| Vitamin D dependent Rickets | 1 (2.2) |
| Brain MRI (abnormal head shape/Chiari malformation, cerebral palsy, cortical blindness, decreased unilateral movement, eye problems, hemangioma (8), hemangioma/possible PHACES syndrome, seizures (2), hand preference, hemiparesis, leukodystrophy, nystagmus (2), sensorineural hearing loss, septal optic dysplasia, subgaleal hematoma) | 19 (41.3) |
| MRI (brachial plexus injury, congenital heart defect, imperforate anus, lymphatic drainage, sacral dimple, spina bifida, spinal cyst/hip dysplasia) | 7 (15.2) |

**Supplemental Table 2: Comparison of Study and Surround Racial and Ethnic Demographics**. Population data for the state of Indiana were obtained by census.gov/quickfacts/IN, which represented population estimates for July 1, 2021.

| Race and Hispanic Origin | Study Population | State of Indiana |
| --- | --- | --- |
| White | 80.4% | 84.2% |
| Black | 6.5% | 10.2% |
| Asian | 2.2% | 2.7% |
| Two or more races | 10.9% | 2.3% |
| Hispanic | 15.2% | 7.7% |

**Supplemental Table 3: Medications Used by Participants.** Four participants were on more than one medication and thus the n is larger than the number of participants.

| Medication | N (%) |
| --- | --- |
| None | 22 (43.1%) |
| Fluconazole | 1 (2.1 %) |
| H2 blocker (Famotidine, Ranitidine) | 3 (5.9%) |
| Imatinib | 1 (2.1 %) |
| Levetiracetam | 2 (3.9%) |
| Levothyroxine | 4 (7.8%) |
| Nitrofurantoin | 1 (2.1 %) |
| Nystatin | 1 (2.1 %) |
| Ondansetron | 1 (2.1 %) |
| Polyethylene glycol | 2 (3.9%) |
| Poly-Vi-Sol | 1 (2.1 %) |
| Poly-Vi-Sol with iron | 2 (3.9%) |
| Timolol | 1 (2.1 %) |
| Vitamin D | 10 (19.6%) |
